# Deep learning-based detection and segmentation of osseous metastatic prostate cancer lesions on computed tomography

**DOI:** 10.1101/2024.11.01.24316594

**Authors:** S J Pawan, Joseph Rich, Shreyas Malewar, Daksh Patel, Matt Muellner, Darryl H Hwang, Xiaomeng Lei, Steven Y Cen, Timothy Triche, Amir Goldkorn, Passant Mohammed, Assad Oberai, Vinay Duddalwar

**Affiliations:** Department of Radiology, Keck School of Medicine, University of Southern California, Los Angeles, CA, USA; Radiomics Lab, Department of Radiology, Keck School of Medicine, University of Southern California, Los Angeles, CA, USA; Department of Biology and Biological Engineering, California Institute of Technology, Pasadena, CA, USA; Center for Personalized Medicine, Dept of Pathology and Laboratory medicine, Children’s Hospital, Los Angeles, CA, USA; Department of Medicine and Biochemistry & Molecular Medicine, USC Norris Comprehensive Cancer Center & Keck School of Medicine, Los Angeles, CA, USA; Department of Aerospace and Mechanical Engineering, Viterbi School of Engineering of the University of Southern California, Los Angeles, CA, USA; Department of Biomedical Engineering, Viterbi School of Engineering of the University of Southern California, Los Angeles, CA, USA

**Keywords:** Computed Tomography, Deep Learning, Image Segmentation, Metastatic Prostate Cancer

## Abstract

**Purpose:** Prostate adenocarcinoma frequently metastasizes to bone and is detected via computed tomography (CT) scans. Accurate detection and segmentation of these lesions are critical for diagnosis, prognosis, and monitoring. This study aims to automate lesion detection and segmentation using deep learning models.

**Methods and Materials:** We evaluated several deep learning models for lesion detection (EfficientNet, ResNet34, DenseNet) and segmentation (nnUNetv2, UNet, ResUNet, ResAttUNet). Performance metrics included F1 score, precision, recall, Area Under the Curve (AUC), and Dice Similarity Coefficient (DSC). Pairwise t-tests compared segmentation accuracy. Additionally, we conducted radiomic analyses to compare lesions segmented by deep learning to manual segmentations

**Results:** EfficientNet achieved the highest detection performance, with an F1 score of 0.82, precision of 0.88, recall of 0.79, and AUC of 0.71. Among segmentation models, nnUNetv2 performed best, achieving a DSC of 0.74, with precision and recall values of 0.73 and 0.83, respectively. Pairwise t-tests showed that nnUNetv2 outperformed ResAttUNet, ResUNet, and UNet in segmentation accuracy (p < 0.01). Clinically, nnUNetv2 also demonstrated superior specificity for lesion detection (0.9) compared to the other models. All models performed similarly in distinguishing diffuse and focal lesions, predicting weight-bearing lesions, and identifying lesion locations, although nnUNetv2 had higher specificity for these tasks. Sensitivity was highest for rib lesions and lowest for spine lesions across all models.

**Conclusions:** EfficientNet and nnUNetv2 were the top-performing models for detection and segmentation, respectively. The radiomic features derived from deep learning-based segmentations were comparable to those from manual segmentations, supporting the clinical applicability of these methods. Further analysis of lesion detection and spatial distribution, as well as lesion quality differentiation, underscores the models’ potential for improving diagnostic workflows and patient outcomes in clinical settings.

## Introduction

Prostate cancer is the most common non-dermatologic cancer in men, and it remains a leading cause of cancer-related mortality^1,2^. The disease often metastasizes to the bones, particularly the spine, pelvis, and ribs, leading to significant morbidity. These lesions often display an osteoblastic appearance. Early and accurate detection of osseous metastatic lesions is critical for effective management and treatment planning. Computed Tomography (CT) is a widely used imaging modality for detecting these metastases due to its high resolution and ability to provide detailed anatomical information^3^ (Fig 1). However, manual interpretation of CT scans can be time-consuming and prone to inter-observer variability, highlighting the need for automated and reliable detection methods. Furthermore, manual interpretation cannot provide quantitative assessment such as radiomics, which must rely on segmentation. Automatically segmenting lesions from multiple CT images with complex background anatomical structures is challenging, as it involves identifying the slice containing the metastatic lesion and then applying the auto-segmentation algorithm.

**Fig 1.**
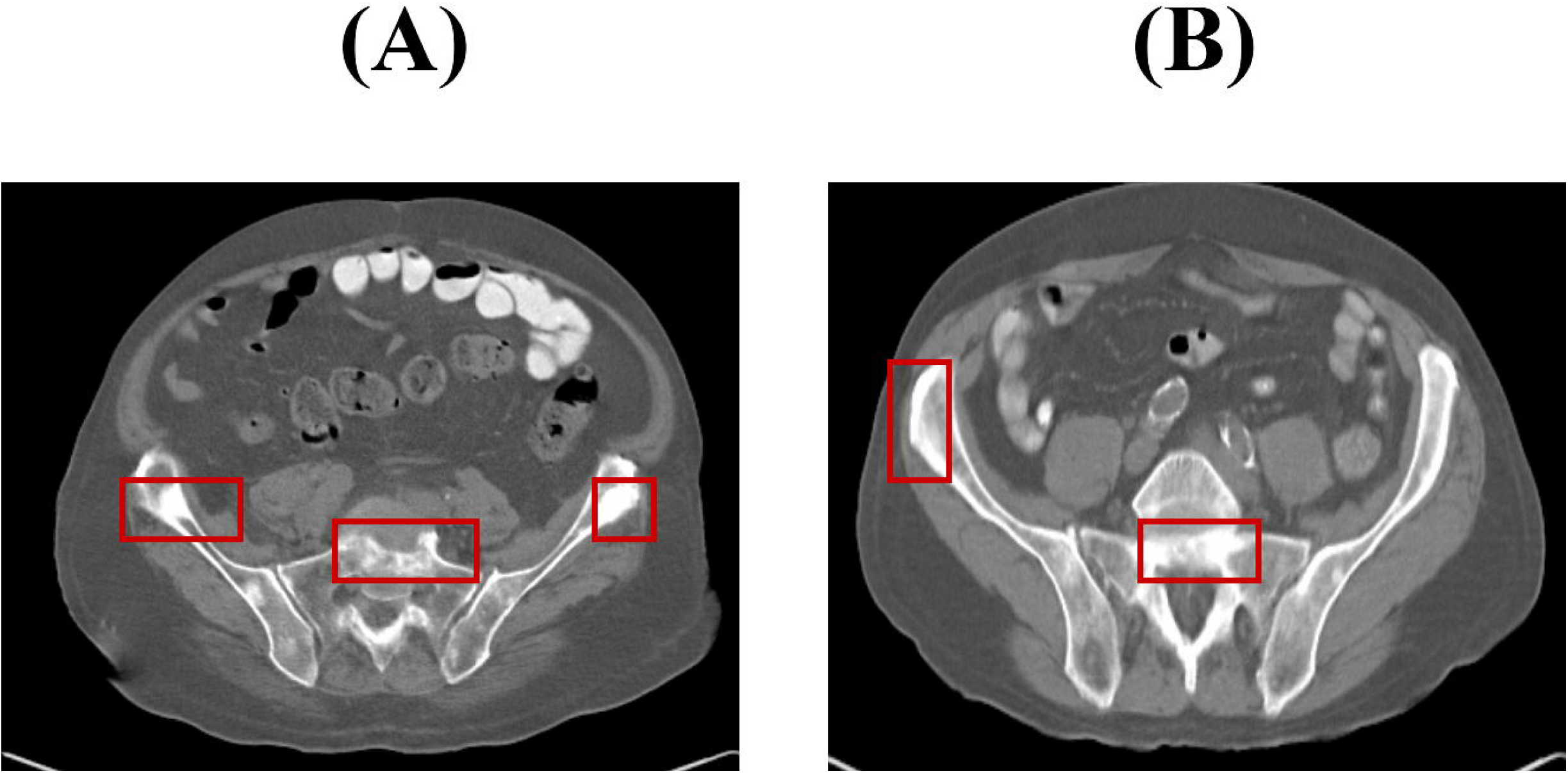
CT scans depicting diffuse osteoblastic metastasis in the bones of the pelvis (highlighted within the bounding box). Broadly, osteoblastic metastatic disease can be identified by increased intensity relative to surrounding tissue and typical bone intensity.

Deep learning has revolutionized medical image analysis, providing powerful tools for the automated classification and segmentation of various pathologies. Convolutional Neural Networks (CNNs) have been extensively utilized for these tasks, significantly enhancing accuracy and efficiency compared to traditional image analysis methods. For detection, deep learning models such as ResNet^4^, DenseNet^5^ and EfficientNet^6^ excel at identifying complex patterns and features within medical images. Similarly, for segmentation, the U-Net^7^, nnUNetv2^8^ characterized by its encoder-decoder structure with skip connections, allows for precise localization and delineation of lesions. These architectures have demonstrated promise in various radiologic analysis tasks, including lesion classification, lesion detection, whole bone and lesion segmentation, and treatment planning ^9-14^. Several studies have explored the application of deep learning for detecting and segmenting osseous metastases in prostate cancer patients.

Chmelik et al.^15^ developed a deep convolutional neural network (CNN) to segment and classify lytic and sclerotic metastatic spinal lesions on 3D CT data, addressing the challenges of ill-defined lesions through automatic feature extraction and incorporating a medial axis transform and Random Forest-based meta-analysis. Moreau et al.^16^ implemented two U-Net based methods to segment bones and bone lesions on 18FDG PET/CT images in metastatic breast cancer, showing improved precision and Dice Similarity Coefficient (DSC) by incorporating bone information into the training process and introducing an automatic PET bone index. Noguchi et al.^17^ created a CNN based on the U-Net architecture for whole-body CT bone segmentation, achieving high accuracy through novel data augmentation techniques and demonstrating generalizability across various datasets. Afnouch et al.^9^ introduced the BM-Seg dataset for bone metastases segmentation and developed the Hybrid-AttUnet++ architecture with dual decoders and an ensemble approach, achieving superior performance compared to existing methods. Chang et al.^18^ developed a deep CNN to detect sclerotic spinal metastases on body CTs, achieving high DSC scores and sensitivity, which demonstrated the potential of deep learning models in detecting sclerotic spinal lesions. Despite the promise of these projects, the task of lesion segmentation for osseous metastatic prostate cancer remains unsolved. This project aims to develop a robust and efficient deep learning-based pipeline for detecting, segmenting, and characterizing osseous metastatic prostate cancer lesions on CT images. By leveraging advanced architectures such as ResNet, DenseNet, Efficient Net for detection and UNet, ResUNet, ResAttUNet, and nnUNetv2 for segmentation, this study aims to achieve high accuracy and reliability in identifying and delineating bone lesions. The development of such a pipeline will eventually enhance diagnostic accuracy and streamline the workflow for radiologists, ultimately contributing to better patient outcomes.

## Materials and Methods

Our IRB-approved, and HIPAA-compliant cohort consists of 8863 2D images across 23 male patients identified with metastatic prostate carcinoma and enrolled in a clinical trial at our institution. Under the supervision of an expert radiologist with 20+ years of experience in radiology and oncologic imaging, the manual binary segmentation of metastatic lesions is carried out to serve as the ground truth. The ground truth mask with pixel-level annotation depicts the lesion and the background. For the detection task, these masks were converted to a binary label corresponding to each image, with 0 indicating the absence of a lesion and 1 indicating the presence of a lesion. We formed an 80:20 ratio to train and test the performance of detection and segmentation methods.

In this study, we utilized three different deep learning architectures for lesion detection, namely ResNet34, DenseNet, and EfficientNet and four different architectures for segmentation, namely UNet, ResUNet, ResAttUNet, and nnUNetv2, respectively. We chose these architectures since they have achieved high accuracy, robustness, and efficiency in various medical imaging tasks. ResNet (Residual Network) utilizes skip connections to enable the training of deep networks by addressing the vanishing gradient problem, enhancing learning efficiency. DenseNet (Dense Convolutional Network) creates direct connections between each layer and all subsequent layers, promoting feature reuse and strengthening gradient propagation. EfficientNet employs a compound scaling method to uniformly scale network depth, width, and resolution, optimizing performance while maintaining computational efficiency. Furthermore, in all the detection models, we have incorporated Gradient-weighted Class Activation Mapping (GradCAM) to ensure the model focuses on the region of interest (ROI) (Fig 2). GradCAM offers visual explanation by highlighting the regions that contribute most significantly to the decision-making process. This technique enhances the interpretability of our models, allowing us to verify that the models accurately identify the ROI during lesion detection. Fig 3 presents a detailed workflow for lesion detection and segmentation of osseous metastatic prostate cancer, followed by the evaluation process incorporating clinical and radiomic analyses.

**Fig 2.**
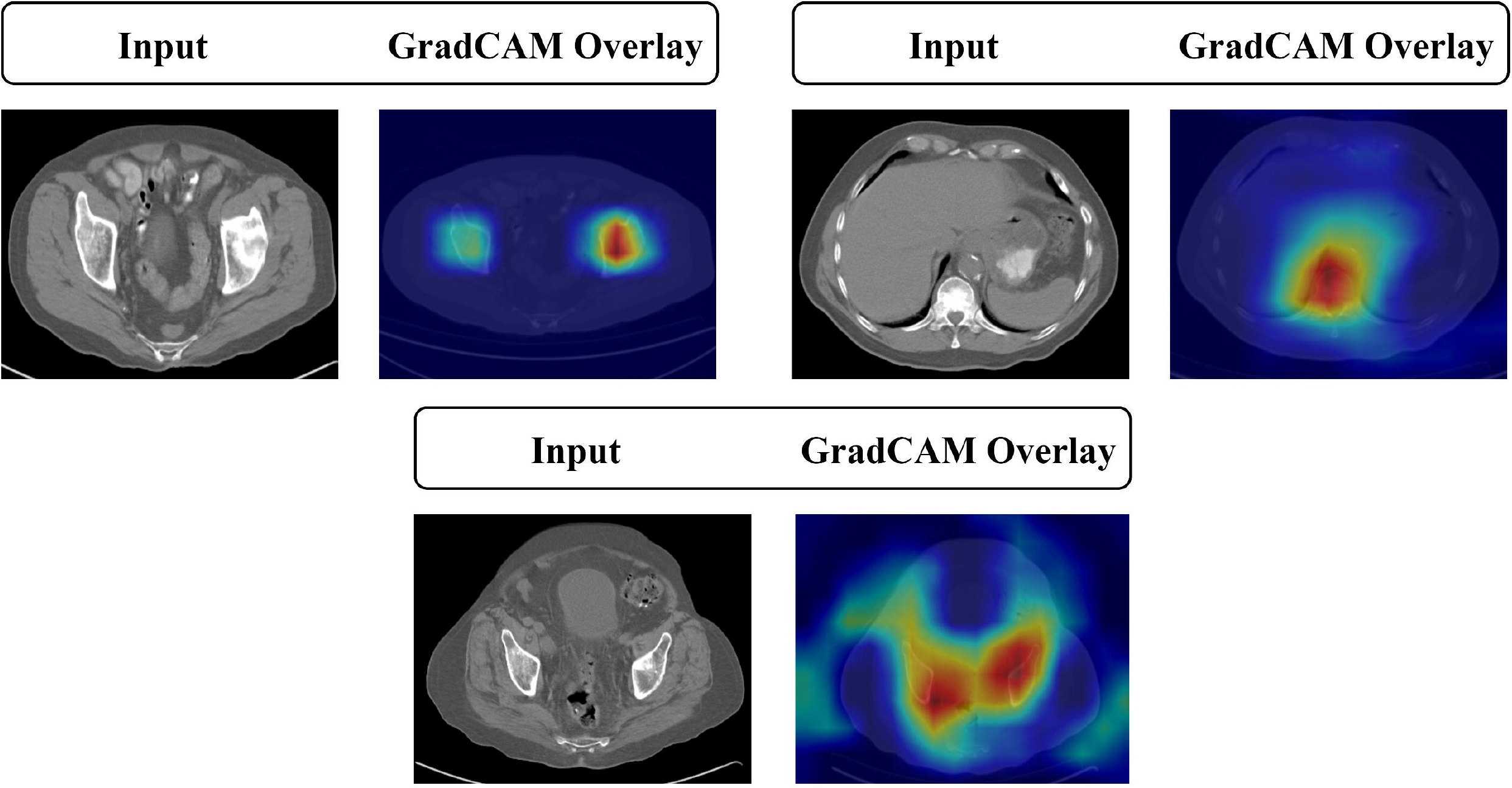
This figure illustrates the regions of the input image that are most influential in the CNN’s decision-making process. The heatmap overlay highlights areas with the highest gradient-based class activation, providing insights into the model’s attention and interpretability of its predictions. Brighter colors indicate regions with higher importance in the final classification, while darker colors signify lesser importance.

**Fig 3.**
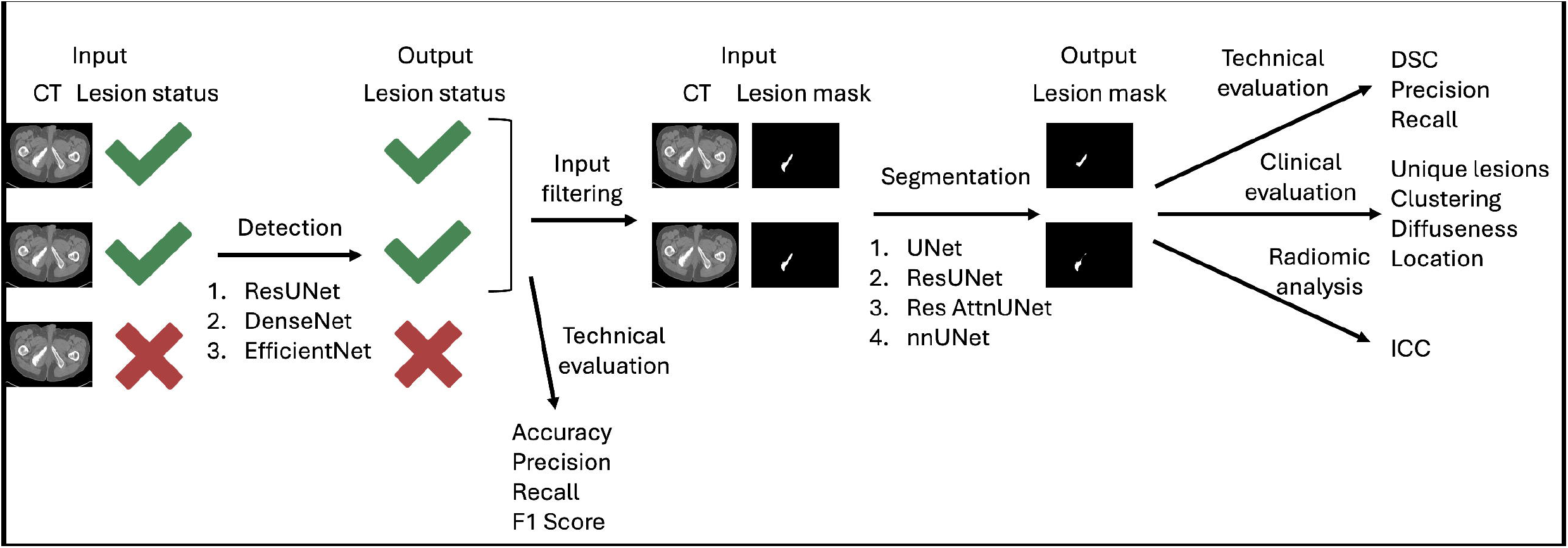
This figure illustrates a comprehensive workflow for lesion detection, segmentation, and evaluation of osseous metastatic prostate cancer lesions. The process begins with lesion detection, where CT images and their lesion status (presence or absence) are processed using detection methods. Following this, the segmentation stage to generate lesion masks. Clinical evaluations are carried out based on unique lesions, clustering, diffuseness, and location, along with radiomic analysis to demonstrate that the radiomic features derived from the lesions segmented by deep learning methods are comparable to manual annotations.

For segmentation tasks, the UNet architecture, known for its encoder-decoder structure with skip connections, excels in precise localization, making it highly effective for biomedical segmentation tasks. ResUNet (Residual Network) is a variant of the UNet architecture that incorporates residual learning, enhancing feature extraction, reuse, and propagation capabilities for more robust performance. The ResAttUNet combines the strengths of UNet and ResUNet, integrating attention mechanisms to focus on relevant parts of the input image, enhancing the network’s ability to model complex dependencies for detailed and accurate segmentation. Lastly, we employed the nnUNetv2 framework, an automated and self-configuring network that adapts to the specific dataset, utilizing a 5-fold cross-validation strategy in the training phase and generating an ensemble of the results to achieve optimal performance. The nnUNetv2 framework offers several key benefits, including its automated and self-configuring design, which adapts to the specific dataset at hand, eliminating the need for manual tuning. Additionally, its extensive data augmentation and preprocessing capabilities streamline the workflow, ensuring consistency and reproducibility in segmentation tasks. By comparing these architectures, we aim to evaluate various state-of-the-art detection and segmentation models comprehensively.

Next, we describe the metric used to quantify the performance of the detection and segmentation algorithms. True positive (TP) refers to a slice (or pixel/voxel) which contains a lesion in both the ground truth (GT) and the prediction. False positive (FP) refers to a slice (or pixel) which contains a lesion in the prediction but not in the ground truth. False negative (FN) refers to a slice (or pixel) which contains a lesion in the ground truth but not in the prediction. True negative (TN) refers to a slice (or pixel) which does not possess a lesion in both the ground truth and the prediction. For the detection task, evaluation metrics include accuracy, precision, recall, F1-score, and AUC. For the segmentation task, evaluation metrics include dice similarity coefficient (DSC), precision, and recall at pixel level. We performed analysis on clinically relevant features on the prediction masks generated by the segmentation models. The clinical analysis tasks are defined as follows:

- Number of 3D lesions detected: Count the number of unique lesions in 3D space, where a single lesion is considered to be a region of positive values in the segmentation mask that are adjacent in 2D space (including diagonal neighbors) or that overlap in at least one neighboring pixel to adjacent slices in 3D space
- Number of spatially clustered lesions: For each lesion in 3D space, count the number of neighboring lesions, where a neighboring lesion is defined as one that is found within 10 slices of another lesion
- Diffuse vs. focal nature: For each lesion in 3D space, determine if the lesion is diffuse or focal throughout the bone which it occupies, where a lesion is considered diffuse if the prediction mask occupies the vast majority of the bone in at least one slice
- Bone identity: For each lesion in 3D space, determine in which bone it resides (skull, rib, spine, pelvis, extremity)
- Weight-bearing nature of bone: For each lesion in 3D space, determine whether or not it resides in a weight-bearing bone (legs, pelvis, or spine)

For the ground truth segmentation masks and the predicted masks from each of the models, we assigned each unique lesion a numeric identifier. For each unique lesion, we determined in which method it appeared (GT, UNet, ResNet, ResAttnNet, nnUNetv2), the number of neighboring lesions, whether the lesion was diffuse or focal (binary), the bone identity of the lesion, and the weight-bearing nature of the bone in which the lesion resides (binary). We conducted a statistical analysis, using the ground truth lesions as anchors for assessing sensitivity and specificity of each model for each clinical task.

## Results

Quantitative results for the detection and segmentation tasks are presented in Fig 4, Table 1, and Table 2, respectively. The detection architectures exhibit more balanced performance, as illustrated in Fig 4a. Among them, EfficientNet achieves the highest performance across all metrics, with an F1 score of 0.82, a precision of 0.88, a recall of 0.79, and an AUC of 0.71. ResNet34 and DenseNet also attain good precision values of 0.87 and 0.85, respectively; however, their recall and AUC values are ssubstantially lower. Among the segmentation models, nnUNetv2 performs the best, achieving a Dice Similarity Coefficient (DSC) of 0.73, along with precision and recall values of 0.73 and 0.83, respectively (Fig 4b). In contrast, the Dice score for the other architectures are appreciably lower. To further quantify these differences in segmentation accuracy, we conducted pairwise paired t-tests across UNet, ResUNet, ResAttUNet and nnUNetv2, using Dice similarity coefficients. ResAttUNet achieved a Dice coefficient of 0.60 ± 0.28, outperforming U-Net, which scored 0.56 ± 0.24, with a mean paired difference of 0.16 ± 0.14 (p < 0.01). Similarly, ResUNet, with a Dice coefficient of 0.58 ± 0.27, slightly outperformed U-Net, which had a Dice coefficient of 0.56 ± 0.24, with a mean paired difference of 0.02 ± 0.13 (p < 0.01). nnUNetv2 model demonstrated superior segmentation accuracy, achieving a Dice coefficient of 0.74 ± 0.21, outperforming ResAttUNet by 0.12 ± 0.37 (p < 0.01), ResUNet by 0.12 ± 0.35 (p < 0.01), and U-Net by 0.16 ± 0.331 (p < 0.01). Supplementary Fig. 1 presents a qualitative analysis of different segmentation methods on slices selected from the test set. The first and second columns display the input and corresponding ground truth, followed by predictions from the remaining methods. These results further illustrate the superior performance of the nnUNetv2 model.

**Table 1.**
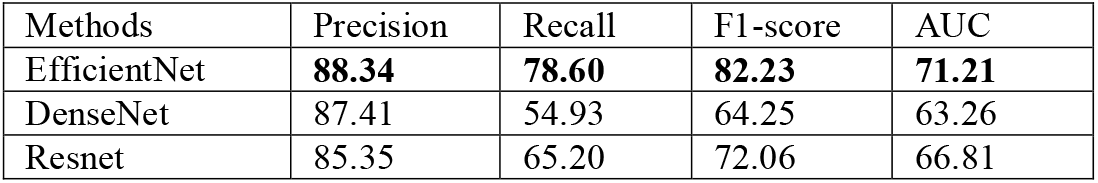
Quantitative analysis of Detection Performance.

**Table 2.** Quantitative Analysis of Segmentation Performance.

| Methods | Dice | Precision | Recall | Parameters<br>(Million) |
| --- | --- | --- | --- | --- |
| Unet | 55.89 | 57.10 | 68.63 | 6.7M |
| ResUNet | 58.97 | 63.86 | 64.27 | 7.7M |
| ResAttUNet | 61.65 | 62.20 | 72.62 | 8M |
| nnUNetv2 | **73.37** | **73.56** | **83.22** | 30.6M |

**Fig 4.**
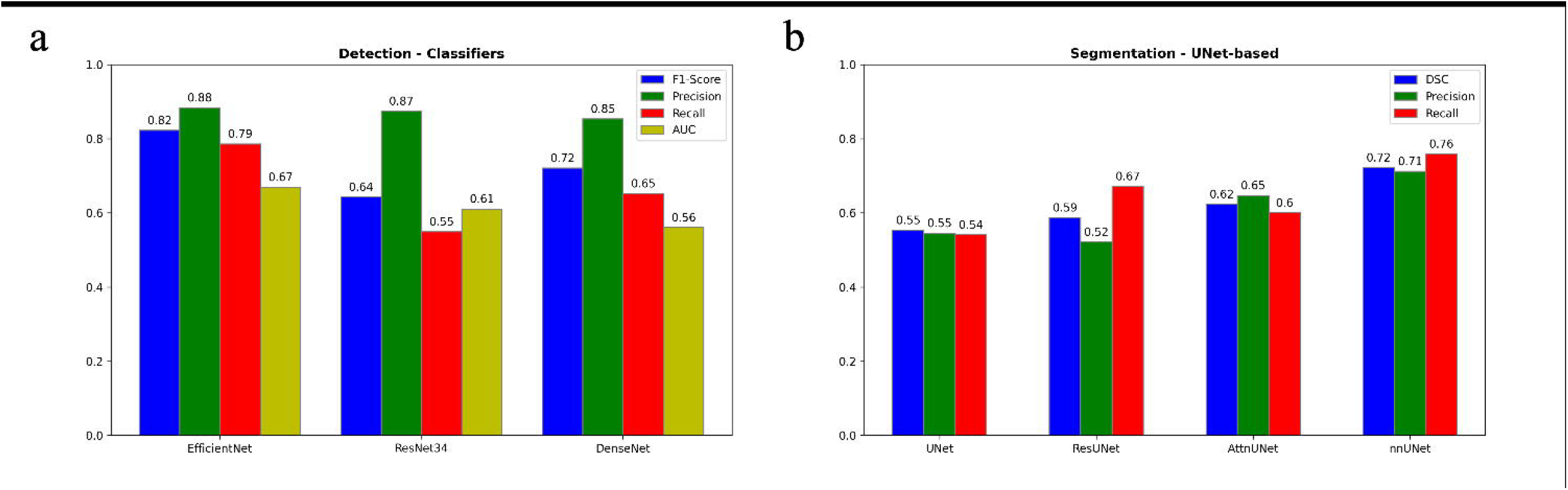
Quantitative detection and segmentation metrics. (a) Detection performance of classifier architectures including EfficientNet, ResNet34, and DenseNet. Blue, F1-Score; green, precision; red, recall; yellow; AUC. (b) UNet-based architectures for the lesion segmentation task. Blue, DSC; green, precision; red, recall.

Further, we interpret the clinical significance of segmentation results through the detection of distinct lesions, the differentiation between focal and diffuse lesion quality, the spatial clustering of lesions, the distribution of predicted lesions across bone locations, and the weight-bearing nature of these locations. Analyzing results based on clinical relevance is crucial because it ensures that the model’s performance aligns with real-world diagnostic needs, enhancing its utility in clinical decision-making and patient outcomes. Although all models exhibited similar sensitivity for lesion detection, nnUNetv2 demonstrated significantly higher specificity at 0.9, compared to 0.48 for UNet, 0.65 for Attention UNet, and 0.46 for ResAttUNet (Fig 5a). All models excelled at distinguishing between diffuse and focal lesions, achieving perfect sensitivity of 1.0 and specificities above 0.9 (Fig 5b). In terms of spatial clustering, UNet predicted the largest spatial clusters, possessing both the highest correct and incorrect counts; nnUNetv2 predicted the smallest spatial clusters, possessing the lowest correct and incorrect counts (Fig 5c). All models showed similar sensitivity for predicting the number of weight-bearing lesions, ranging from 0.3 to 0.5, but nnUNetv2 had the highest specificity at 0.95, compared to 0.6 to 0.8 for the other methods (Fig 5d). When predicting lesion locations, all models had similar sensitivity and high specificity (∼1), but with lower sensitivity overall (Fig 5e). The models globally had the lowest sensitivities for predicting spine lesions, with values between 0.25-0.4, and the highest sensitivities for predicting rib lesions, with values between 0.6-1.0 (Fig 5e).

**Fig 5.**
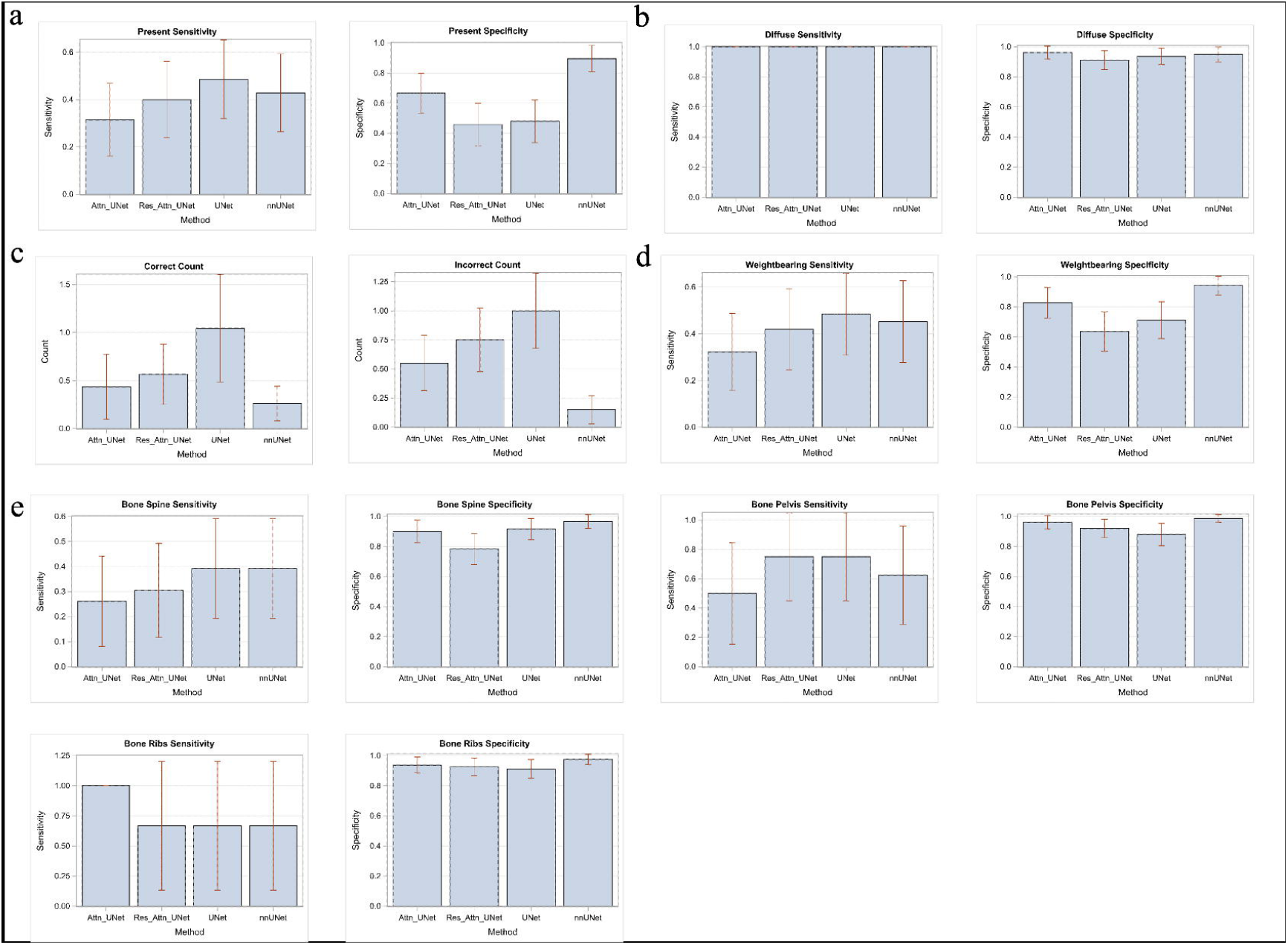
Clinical segmentation metrics, with sensitivity and specificity relative to ground truth. (a) Lesion prediction. (b) Diffuse vs. focal lesion status. (c) Local clustering of lesions. (d) Weight-bearing nature of the predicted lesions. (e) Fraction of lesions in each body part – spine, pelvis, and ribs.

Often, segmented ROIs for lesions are used as input to algorithms that evaluate radiomic features. Given this, we wish to determine whether using ROIs generated by the deep learning algorithm versus those determined by the expert radiologist can have an impact on final radiomic features. To evaluate this the Intraclass Correlation Coefficient (ICC) between radiomic features extracted from algorithm-predicted ROIs and those obtained from manually segmented ROIs was evaluated. Across all metrics the nnUNetv2 segmentation has the largest values of ICC values indicating that it is closest to the manual segmentation (Table 3).

**Table 3.** Comparative Analysis of Intraclass Correlation Coefficient (ICC) Across Various Methods for Radiomic Feature Assessment.

| ICC (Min, Max) |  |  |  |  |  |  |
| --- | --- | --- | --- | --- | --- | --- |
|  | First | GLCM | GLDM | GLRLM | GLSZM | NGTDM |
| UNet | 0.93 (0.92, 0.93) | 0.84 (0.83, 0.85) | 0.92 (0.91, 0.93) | 0.87 (0.86, 0.88) | 0.83 (0.81, 0.84) | 0.73(0.69,0.77) |
| ResUNet | 0.96 (0.95, 0.96) | 0.81 (0.79, 0.82) | 0.89 (0.88, 0.90) | 0.82 (0.80, 0.83) | 0.81 (0.79, 0.83) | 0.40 (0.32, 0.47) |
| ResAttUNet | 0.97 (0.97, 0.98) | 0.88 (0.87, 0.89) | 0.94 (0.94, 0.95) | 0.89 (0.88, 0.90) | 0.89 (0.88, 0.90) | 0.46 (0.39, 0.53) |
| nnUNetv2 | 1.0(1.0,1.0) | 0.91 (0.90, 0.91) | 0.95 (0.95, 0.96) | 0.95 (0.94, 0.95) | 0.94 (0.93, 0.94) | 0.95 (0.94,0.95) |

## Discussion

The findings presented in this study highlight the performance and clinical relevance of various deep learning architectures for the detection and segmentation of lesions. Our results indicate that nnUNetv2 consistently outperforms other models across multiple metrics, both in segmentation accuracy and in preserving the integrity of radiomic features. The detection architectures evaluated in this study demonstrate a balanced performance, with EfficientNet emerging as the most effective model, achieving the highest scores across all metrics, including F1 score, precision, recall, and AUC. This model’s ability to maintain high precision and recall suggests a well-calibrated approach to managing false positives and false negatives, which is crucial in clinical settings where misclassification can have significant consequences. While ResNet34 and DenseNet also perform decently, their lower recall and AUC scores indicate that these models may miss certain lesions, reducing their reliability in practice. Among the segmentation models, nnUNetv2 distinctly outperforms its counterparts, achieving superior Dice Similarity Coefficients (DSC) and demonstrating higher precision and recall. This performance is likely due to nnUNetv2’s robust architecture, which includes extensive data augmentation and advanced normalization techniques that enhance generalization across different datasets. The pairwise comparisons further validate nnUNetv2’s superiority, with statistically significant improvements over ResUNet, U-Net, and ResAttnUNet.

The clinical relevance of the segmentation results was further explored by analyzing the models’ ability to detect distinct lesions, differentiate between focal and diffuse lesion qualities, and predict lesion distribution across various bone locations. nnUNetv2 not only demonstrated higher overall accuracy but also excelled in specificity, particularly in distinguishing between different types of lesions and in predicting the spatial clustering of lesions. This is a critical aspect in clinical setting, where the accurate classification of lesion types and their spatial distribution can directly influence treatment decisions. The higher specificity observed with nnUNetv2, especially in weight-bearing lesions, underscores its potential utility in clinical workflows, where precision in identifying critical lesion locations can be pivotal.

Radiomics enhances diagnostic and prognostic capabilities by extracting quantitative features from medical imaging, but its effectiveness relies on accurate initial segmentation. We analyzed the radiomic features extracted from segmentations predicted by different methods and compared them to those derived from ground truth (GT) segmentations. We evaluated how effectively each deep learning architecture preserved these radiomic features compared to the GT. Our study reveals significant variability in the Intraclass Correlation Coefficient (ICC) values across different segmentation models and radiomic features. The lower ICC values associated with NGTDM across all models, except nnUNetv2, suggest that this texture feature is particularly sensitive to misalignment in the region of interest (ROI). nnUNetv2’s superior performance in maintaining high ICC values across a broad range of radiomic features, including NGTDM, indicates that it offers the most reliable segmentation output for radiomics. The narrow confidence intervals observed in the ICC values for nnUNetv2 further emphasize its consistency and robustness, making it a preferred choice for studies where radiomic analysis is integral. In contrast, the lower ICC values observed in ResUNet and ResAttnUNet indicate potential limitations in their application for radiomic feature extraction.

While this study provides valuable insights into the performance of different segmentation models, some limitations need to be acknowledged. The dataset used for training and evaluation may not encompass the full variability encountered in clinical practice, particularly regarding lesion types, sizes, and locations. In subsequent studies, we aim to include more diverse cases, which would enhance the generalizability of the findings. While nnUNetv2 demonstrated superior performance in this study, it is important to consider the computational demands associated with its training and deployment. The same holds true for the EfficientNet, which performed well in the detection task. The high computational cost may limit its accessibility in resource-constrained environments. Developing more efficient versions of nnUNetv2 or alternative models that offer a better balance between performance and computational requirements will be explored. Finally, the integration of radiomics with segmentation models, as demonstrated in this study, highlights the potential for personalized medicine. However, the clinical translation of these findings will require rigorous validation through large-scale, multi-center studies.

## Data Availability Statement

Research data are not available at this time.

## Acknowledgements

We would like to thank the rest of the Radiomics Lab at USC for their feedback.

