## Supplementary figures and images for "Deep learning-based detection and segmentation of osseous metastatic prostate cancer lesions on computed tomography"

### Supplemental Figure 1

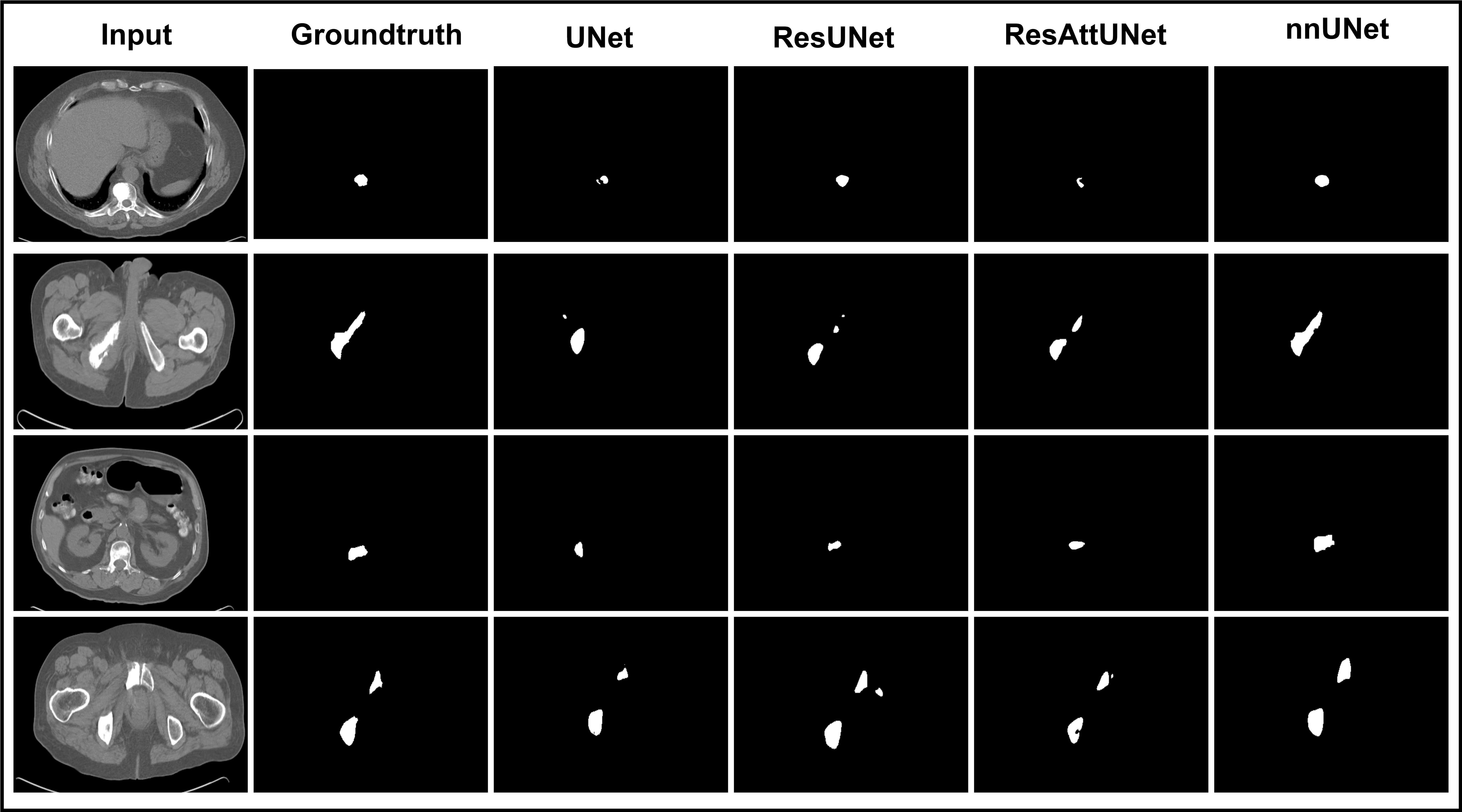
